# Association between Clinical, Laboratory and CT Characteristics and RT-PCR Results in the Follow-up of COVID-19 patients

**DOI:** 10.1101/2020.03.19.20038315

**Authors:** Hang Fu, Huayan Xu, Na Zhang, Hong Xu, Zhenlin Li, Huizhu Chen, Rong Xu, Ran Sun, Lingyi Wen, Linjun Xie, Hui Liu, Kun Zhang, Chuan Fu, Keke Hou, Zhigang Yang, Ming Yang, Yingkun Guo

**Author notes:** Guarantor and correspondent: Ying-kun Guo, MD, Department of Radiology; Key Laboratory of Obstetric & Gynecologic and Pediatric, Diseases and Birth Defects of Ministry of Education; West China Second University Hospital, Sichuan University; 20# South Renmin Road, Chengdu, Sichuan 610041, China. Hang Fu, Huayan Xu, Na Zhang, are co-authors. Yingkun Guo, Ming Yang, Zhigang Yang and Keke Hou contributed equally to this work and should be considered as co-corresponding authors. **Conflict of Interest: none declared**.

## Abstract

**Background:** Since December 2019, more than 100,000 coronavirus disease 2019 (COVID-19) patients have been confirmed globally based on positive viral nucleic acids with real-time reverse transcriptase-polymerase chain reaction (RT-PCR). However, the association between clinical, laboratory and CT characteristics and RT-PCR results is still unclear. We sought to examine this association in detail, especially in recovered patients.

**Methods:** We analysed data from 52 confirmed patients who had been discharged with COVID-19. The clinical, laboratory, and radiological data were dynamically recorded and compared with the admission and follow-up RT-PCR results.

**Results:** In this cohort, 52 admitted COVID-19 patients who had confirmed positive RT-PCR results were discharged after 2 rounds of consecutively negative RT-PCR results. Compared with admission levels, CRP levels (median 4.93 mg/L [IQR: 1.78-10.20]) decreased significantly (p<0.001). and lymphocyte counts (median 1.50×109/L [IQR: 1.11-1.88]) increased obviously after obtaining negative RT-PCR results (p<0.001). Additionally, substantially improved inflammatory exudation was observed on chest CT except for 2 progressed patients. At the two-week follow-up after discharge, 7 patients had re-positive RT-PCR results, including the abovementioned 2 progressed patients. Among the 7 patients, new GGO was demonstrated in 2 patients. There were no significant differences in CPR levels or lymphocyte counts when comparing the negative and re-positive PCT results (all p >0.05).

**Conclusion:** Heterogeneity between CT features and RT-PCR results was found in COVID-19, especially in some recovered patients with negative RT-PCR results. Our study highlights that both RT-PCR and chest CT should be considered as the key determinants for the diagnosis and management of COVID-19 patients.

## Introduction

An outbreak of novel coronavirus disease 2019 (COVID-19) emerged in Wuhan, China, and spread quickly across 114 countries around the world. On 11 March 2020, the Word Health Organization(WHO) made the assessment that COVID-19 can be characterized as a pandemic because we have now surpassed 100,000 confirmed cases worldwide, with more than 4,000 deaths[1,2]. According to the recommended guidelines, approximately 22,888 patients were discharged in China after two consecutive negative real-time polymerase chain reaction (RT-PCR) results of SARS-CoV-2 nucleic acid[3,4]. Until now, there has been no adequate, approved, or available alternative to the emergency use of RT-PCR for diagnosing COVID-19. Thus, nucleic acid testing plays an important and key role in the diagnosis and therapeutic decision-making in COVID-19. Apart from the RT-PCR results, new coronavirus pneumonia has been observed in approximately 76.4% of SARS-CoV-2-infected patients on chest CT images[5]. Although the epidemiological, clinical, laboratory and CT characteristics of COVID-19 have been well documented, their association with RT-PCR results is still unclear [5-11]. Recently, follow-up examinations have found that few patients have positive RT-PCR results after recovery[12]. However, the follow-up of recovered patients has been limited to date. We sought to assess the association between RT-PCR results and the clinical, laboratory and radiological features of COVID-19 patients. Specifically, we evaluated the clinical characteristics of patients with re-positive RT-PCT test results after recovery from COVID-19.

## Materials and methods

### Population

For this single-centre study, we retrospectively recruited patients diagnosed with COVID-19 from Jan 1 to Feb 20, 2020, at Chengdu Public Health Clinical Medical Center, which is a hospital that specializes in infectious diseases, and it is also a designated hospital for this emergency in Chengdu, China. Our inclusion criteria were as follows: (1) patients with laboratory-confirmed SARS-CoV-2 infection by RT-PCR results [13] and (2) patients who were discharged from the hospital with standard treatment after having two consecutive negative RT-PCR results based on the sixth guidelines of the CDC in China [4]. Patients who died or were not discharged were excluded. The institutional ethics board of our institutes approved this study (No. 2020.43). All patients were orally informed that their clinical data would be used to perform this research to contribute to the deeper understanding of COVID-19 and that their private information would be strictly confidential. The RT-PCR results after recovery from COVID-19 were tracked, and the due date was Feb 29, 2020.

## CT scan and imaging review

A series of thin-section chest CT scans was performed with a GE-BrightSpeed spiral CT (16-section detector; 120 KV/400 mA; slice thickness of 1.25 mm). Images were acquired under end-inhalation breath-holding conditions. The scanning range covered the area from the angle of the costal diaphragm at the base of the lung to the entrance of the chest. A standard lung window (window level -430-550 HU, window width 1150-1350 HU) and mediastinal window (window level 35-40 HU, window width 350-400 HU) were used for imaging assessments.

Two trained radiologists with at least 4 years of experience in cardiothoracic imaging who were blinded to the clinical information reviewed the CT images of the enrolled subjects independently. A third cardiothoracic radiologist with more than 8 years of experience was references when disagreement was encountered. The series of CT scans was evaluated for the presence of the following characteristics: (1) ground-glass opacities, (2) consolidation, (3) mixed ground-glass opacities, (4) vascular enlargement, (5) interlobular septal thickening, (6) reticulation, (7) number of involved lobes, and (8) progression or improvement of lesions in each lobe.

### Data collection

All medical records were reviewed carefully, and data were collected in standard form. The data we recorded included demographic and epidemic information, symptoms and signs, treatment strategies, complications, underlying comorbidities, and subtypes of 2019 novel coronavirus pneumonia (NCP). Data from the laboratory tests were also recorded, including blood count, CRP level, coagulation, cardiac markers, liver function, renal function, peripheral blood lymphocyte subsets and so on. The date of the negative RT-PCR results was defined as the day of the first of two consecutive negative RT-PCR results. The date of disease onset was defined as the day that symptoms or signs were first identified. NCP was classified into mild, common, severe and critically severe subtypes according to the guidelines of the CDC in China [4].

### Statistical analysis

Continuous variables are presented as the mean ± standard difference (SD) or median and interquartile range (IQR). Categorical variables are presented as frequencies and percentages. Continuous variables were compared using matched group ANOVA and independent t tests (normal distribution) or matched group Friedman and Wilcoxon signed rank tests. P<0.05 was considered statistically significant. All analyses were performed with offline software (SPSS, version 22, International Business Machines, Armonk, New York, USA).

## Results

### Clinical characteristics at admission

A total of 52 laboratory-confirmed COVID-19 patients discharged from the hospital were enrolled. The basic clinical characterization is shown in Table 1. The median age was 44.5 (IQR: 33.0-56.5) years, 11 patients (21.2%) were aged over 60 years, and 28 patients (53.8%) were male. Patients were living in Wuhan (n=13, 25%) or had recently travelled to Wuhan (n=15, 53.8%)), a portion of whom (n=18, 34.6%) had been exposed to COVID-19 patients, and only 6 (11.5%) cases were of unknown origin. The median interval from disease onset to admission was 5 days (IQR: 3-7). Fever was the most common symptom; fatigue, dry cough, expectoration, etc., were also found. More than half of these patients (73.1%) had common NCP, and a few had severe (n=10, 19.2%) and critically severe (n=4, 7.7%) NCP. Patients received antibiotic treatment (n=12, 23.1%) and traditional Chinese medicine (n=32, 61.5%) if necessary. Three (5.8%) patients needed intensive unit care, and 4 (7.7%) patients required non-invasive mechanical ventilation or high-flow oxygen therapy. Three (5.8%) patients developed acute respiratory distress syndrome, and 5 (9.6%) patients had type I respiratory failure.

**Table 1.**
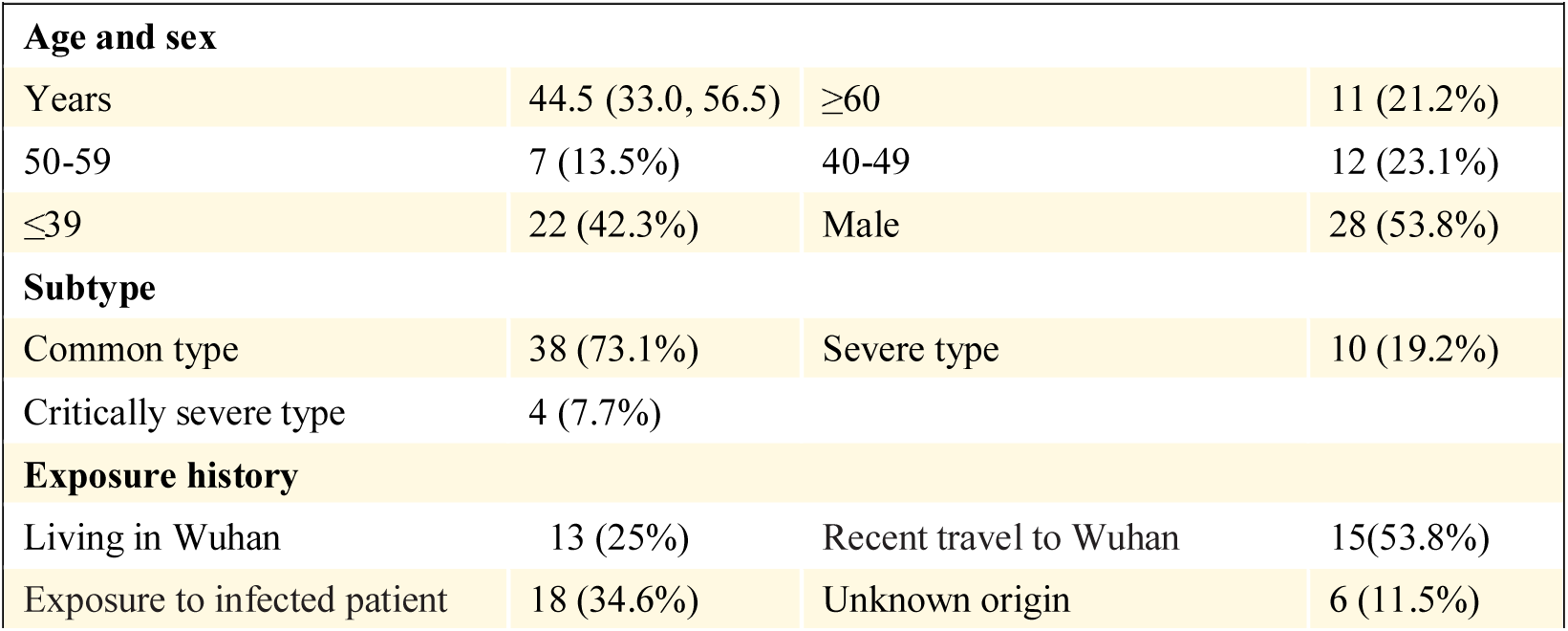
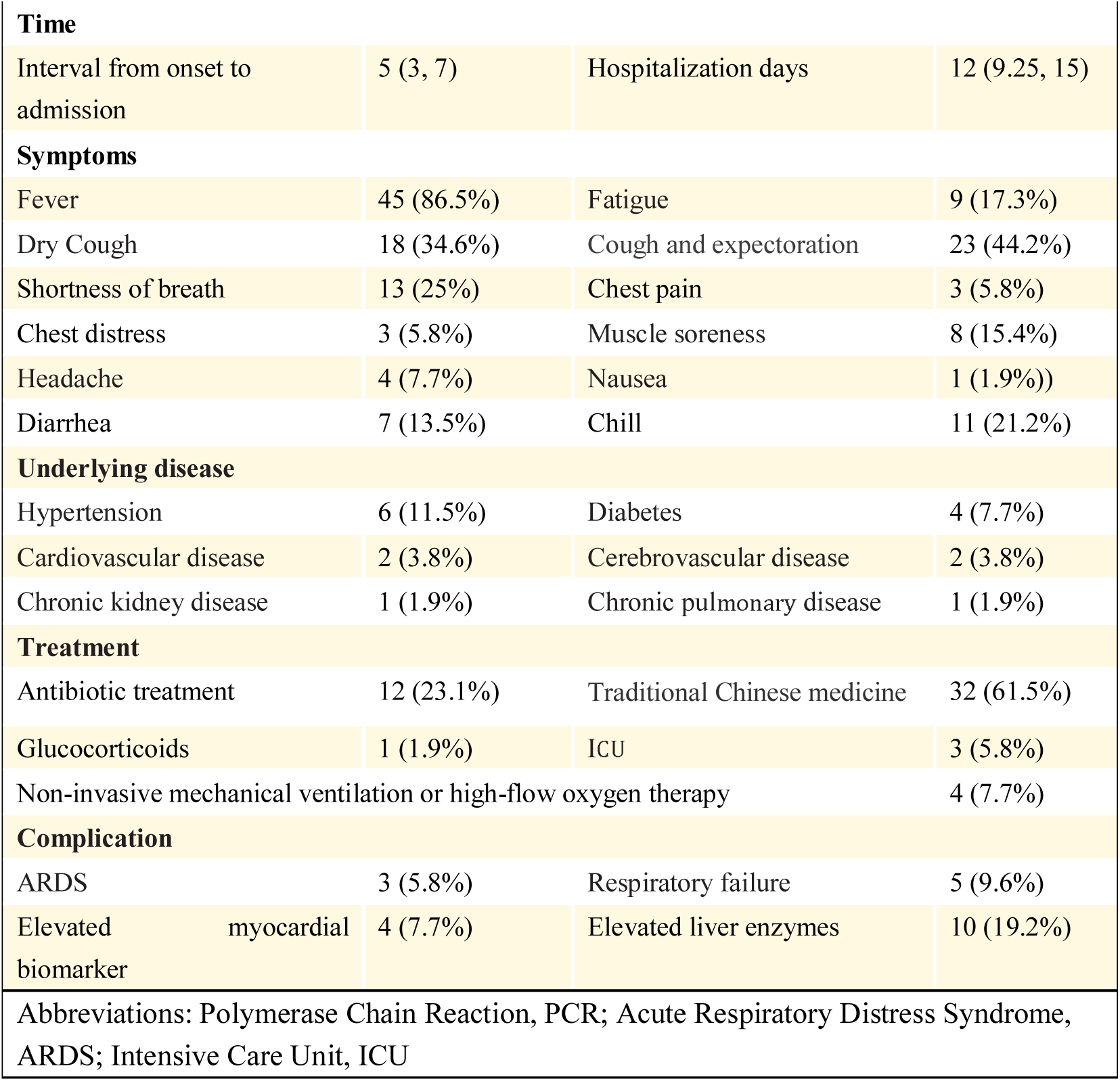
Clinical characteristics at admission (n=52)

The laboratory test results at admission are shown in Table 2. The white blood cell count (n=35, 67.3%) and neutrophil count (n=37, 71.2%) were in the normal range in most patients. Fifteen (28.8%) patients had reduced lymphocyte counts, and the remaining patients had counts within the normal range. In terms of CRP, most patients had increased level at admission except for 19 (36.5%) patients.

**Table 2.**
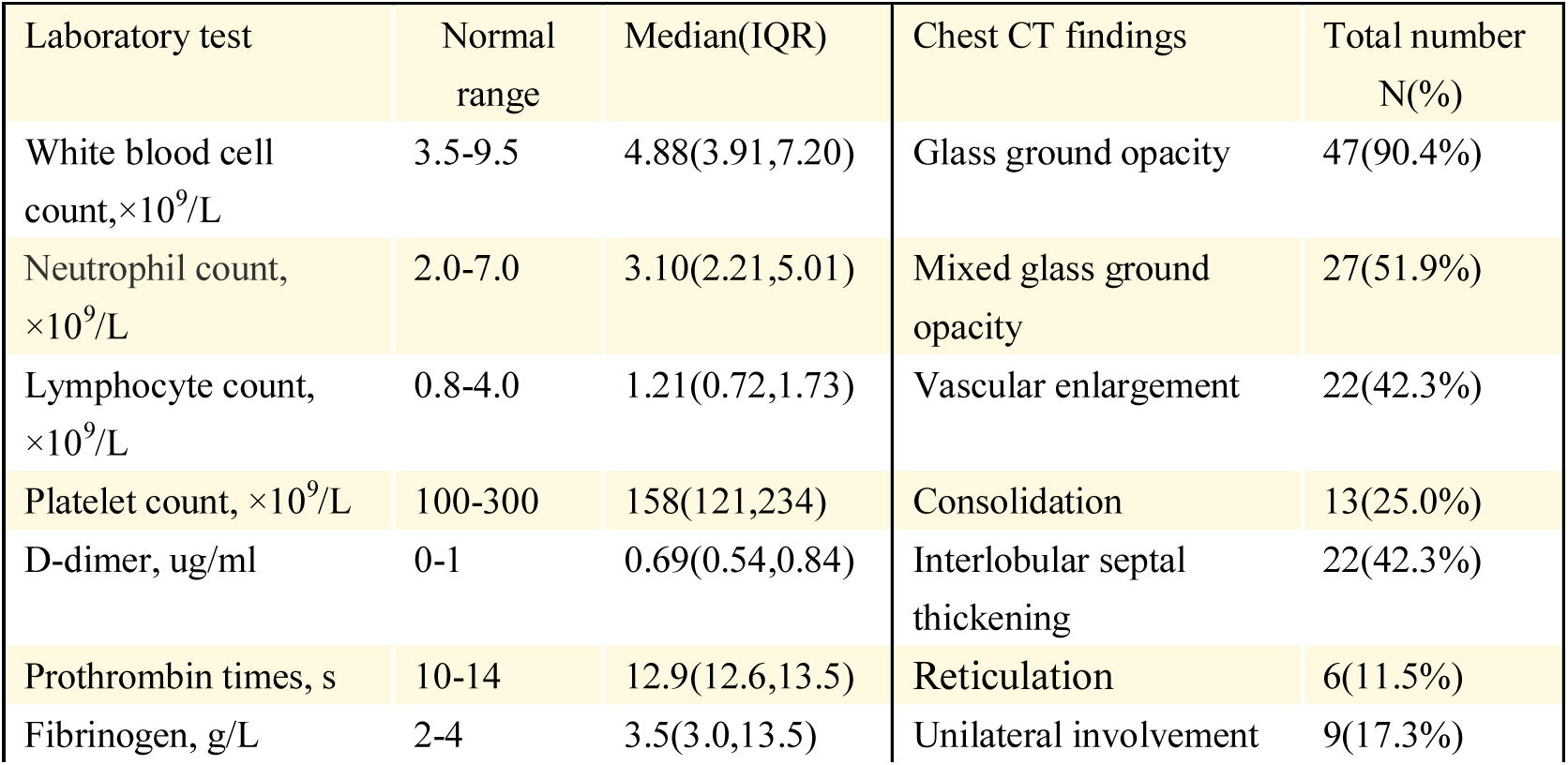
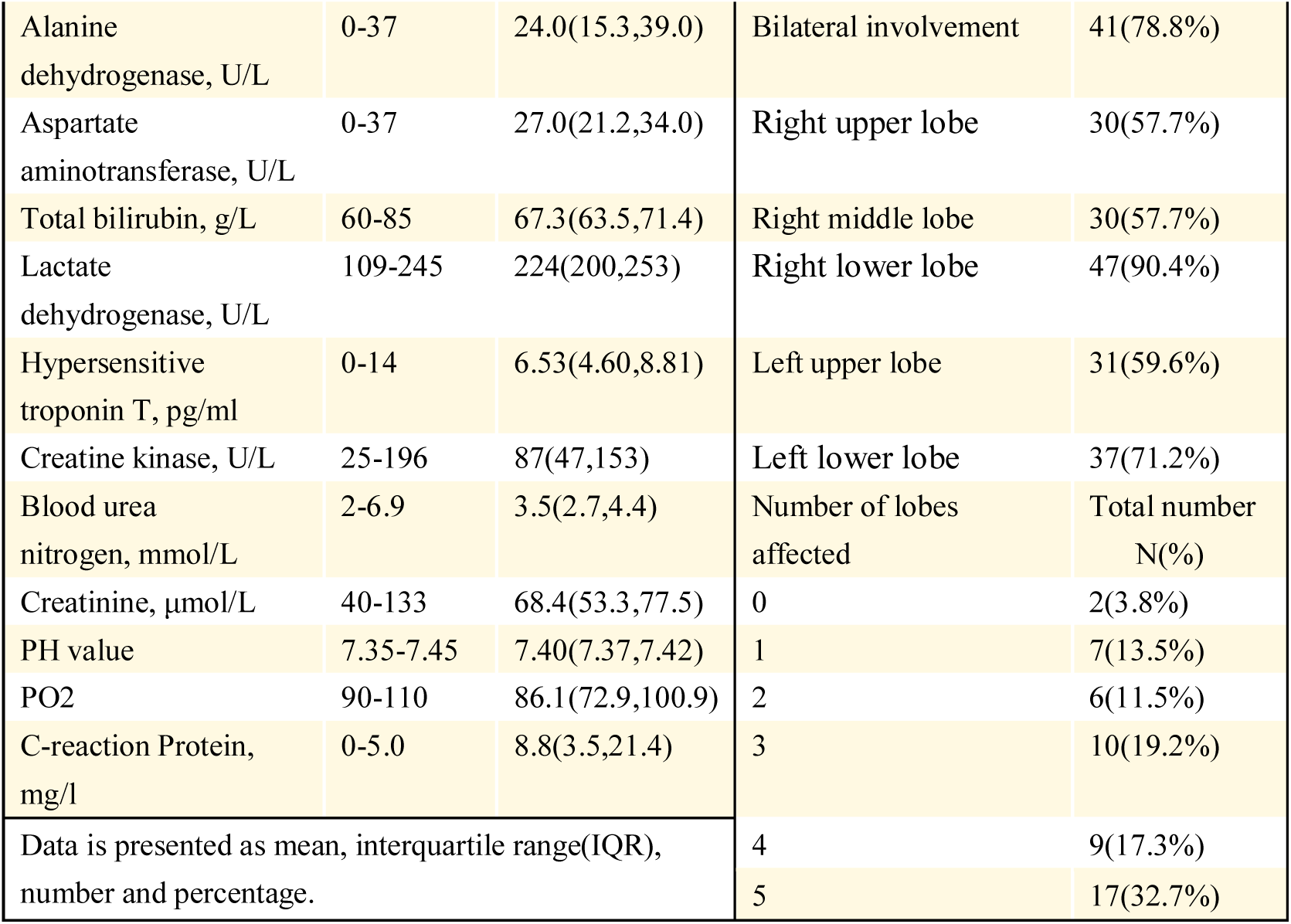
Laboratory test and radiographical signs on chest CT at admission

| Laboratory test | Normal range | Median(IQR) | Chest CT findings | Total number N(%) |
| --- | --- | --- | --- | --- |
| White blood cell count, $\times 10^9/L$ | 3.5-9.5 | 4.88(3.91,7.20) | Glass ground opacity | 47(90.4%) |
| Neutrophil count, $\times 10^9/L$ | 2.0-7.0 | 3.10(2.21,5.01) | Mixed glass ground opacity | 27(51.9%) |
| Lymphocyte count, $\times 10^9/L$ | 0.8-4.0 | 1.21(0.72,1.73) | Vascular enlargement | 22(42.3%) |
| Platelet count, $\times 10^9/L$ | 100-300 | 158(121,234) | Consolidation | 13(25.0%) |
| D-dimer, ug/ml | 0-1 | 0.69(0.54,0.84) | Interlobular septal thickening | 22(42.3%) |
| Prothrombin times, s | 10-14 | 12.9(12.6,13.5) | Reticulation | 6(11.5%) |
| Fibrinogen, g/L | 2-4 | 3.5(3.0,13.5) | Unilateral involvement | 9(17.3%) |
| Alanine dehydrogenase, U/L | 0-37 | 24.0(15.3,39.0) | Bilateral involvement | 41(78.8%) |
| Aspartate aminotransferase, U/L | 0-37 | 27.0(21.2,34.0) | Right upper lobe | 30(57.7%) |
| Total bilirubin, g/L | 60-85 | 67.3(63.5,71.4) | Right middle lobe | 30(57.7%) |
| Lactate dehydrogenase, U/L | 109-245 | 224(200,253) | Right lower lobe | 47(90.4%) |
| Hypersensitive troponin T, pg/ml | 0-14 | 6.53(4.60,8.81) | Left upper lobe | 31(59.6%) |
| Creatine kinase, U/L | 25-196 | 87(47,153) | Left lower lobe | 37(71.2%) |
| Blood urea nitrogen, mmol/L | 2-6.9 | 3.5(2.7,4.4) | Number of lobes affected | Total number<br>N(%) |
| Creatinine, μmol/L | 40-133 | 68.4(53.3,77.5) | 0 | 2(3.8%) |
| PH value | 7.35-7.45 | 7.40(7.37,7.42) | 1 | 7(13.5%) |
| PO2 | 90-110 | 86.1(72.9,100.9) | 2 | 6(11.5%) |
| C-reaction Protein, mg/l | 0-5.0 | 8.8(3.5,21.4) | 3 | 10(19.2%) |
| Data is presented as mean, interquartile range(IQR), number and percentage. |  |  | 4 | 9(17.3%) |
|  |  |  | 5 | 17(32.7%) |

The radiographic signs on admission according to chest CT are shown in Table 2. At the initial CT examination, 2 (3.8%) patients had normal chest CT results, 41 (78.8%) patients had bilateral infection, and 9 (17.3%) patients had unilateral infection. The lower lobes of the lungs had a higher incidence of involvement: 47 (90.4%) cases involved the right lower lobe, and 37 (71.2%) cases involved the left lower lobe. GGO (90.4%) was the most common finding, which was accompanied by other signs, including consolidation (25.0%), mixed GGO (51.9%), interlobular septal thickening (42.3%), vascular enlargement (42.3%), and reticulation (11.5%).

### Follow-up results of laboratory tests before and after negative RT-PCR results

By comparing the laboratory test results at different times (shown in Table 3), we found that most of these indicators did not change significantly (all p>0.05), except for the lymphocyte count, CRP level, and peripheral blood lymphocyte subset. The lymphocyte count (median 1.50×10^9^ /L [IQR: 1.11-1.88]) increased obviously after obtaining negative RT-PCR results (p<0.001). In addition, the CD3+ count, CD3+CD4+ count, and CD3+CD8+ count increased significantly after obtaining negative RT-PCR results (all p<0.001). Furthermore, after obtaining negative RT-PCR results, the CRP level was decreased (9.47 (3.78, 24.20) mg/l vs. 10.25 (2.62, 22.61) mg/l vs. 4.93 (1.78, 10.2) mg/l, p=0.000). Nevertheless, 15 (50.0%) patients still had elevated CRP levels after obtaining negative RT-PCR results.

**Table 3.**
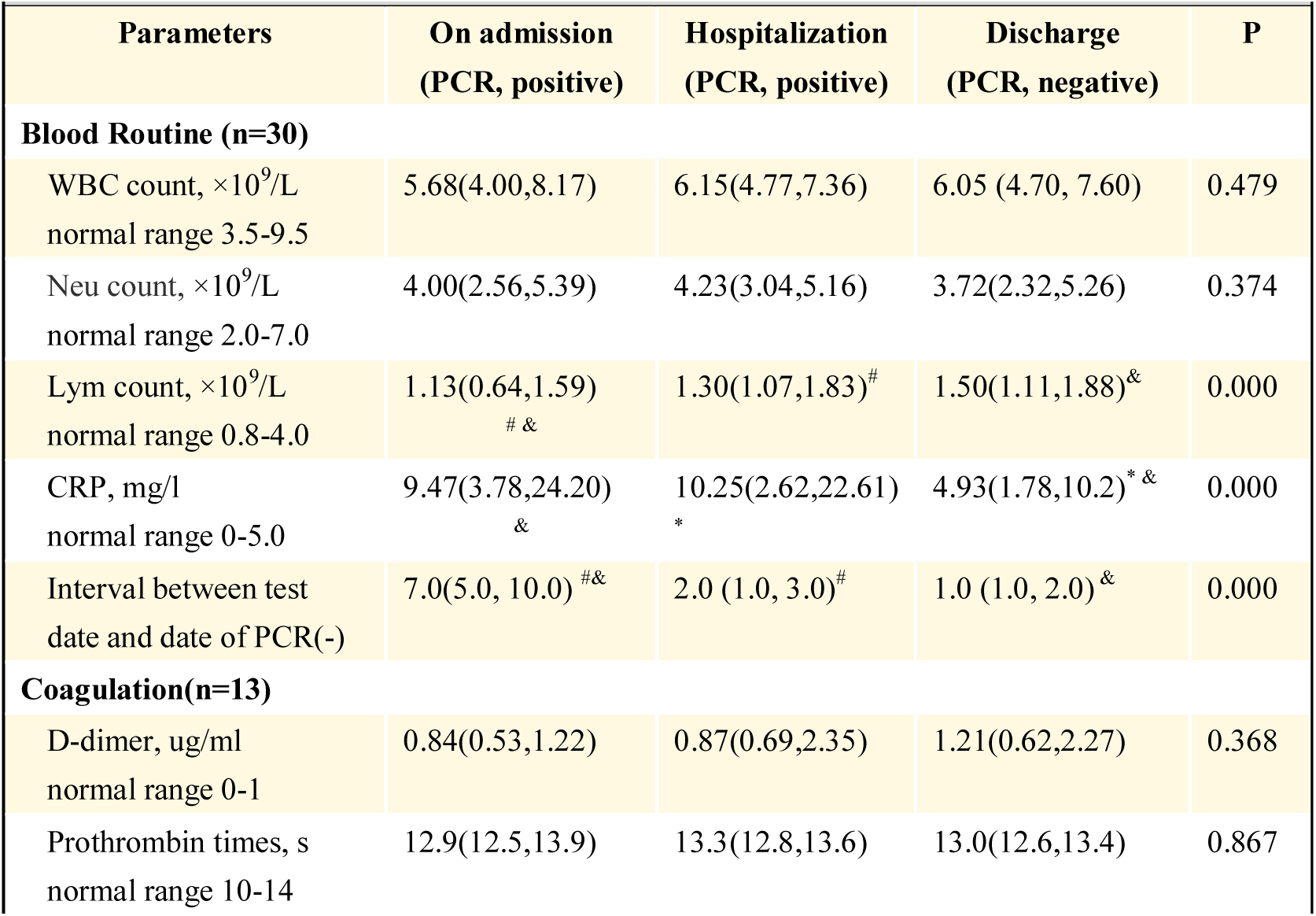
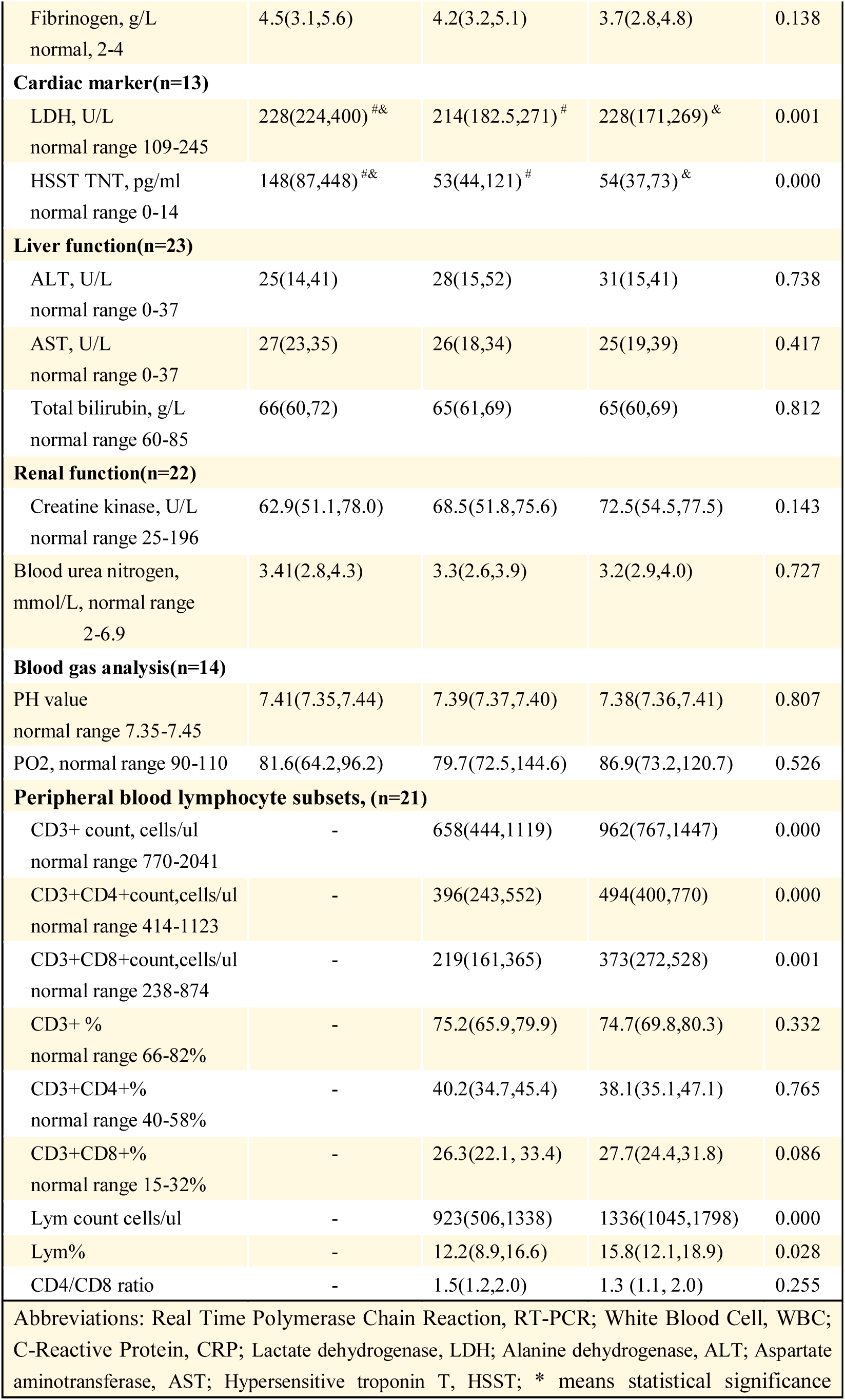
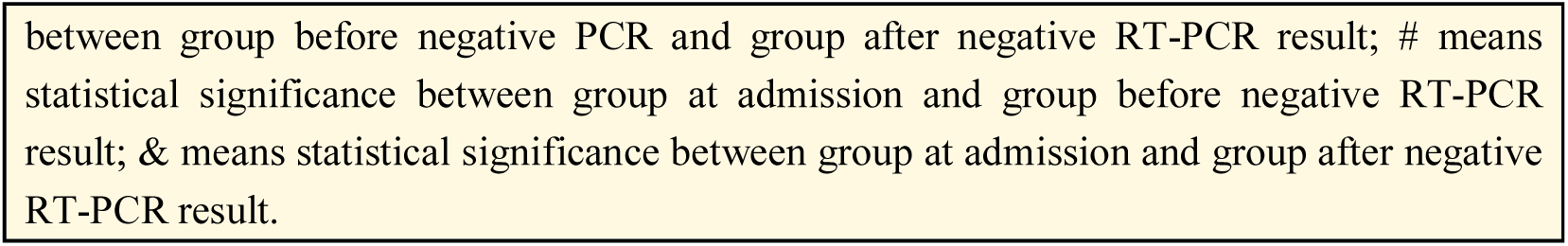
Follow-up of laboratory tests before and after SARS-CoV-2 being negative by RT-PCR result

### Follow-up chest CT results before and after negative RT-PCR results

Among the enrolled subjects, 18 patients underwent chest CT for three times, including exams at admission as well as before and after obtaining negative RT-PCR results. The detailed radiographic signs are listed in Table 4. Compared with the 1^st^ CT results at admission, the 2^nd^ chest CT results with positive RT-PCR results in the hospital demonstrated heterogeneity of lesions in the different lobes of the lung: 1) right upper lobe improved in 4 (22.2%) patients and progressed in 6 (33.3%) patients; 2) right middle lobe improved in 5 (27.8%) patients and progressed in 4 (22.2%) patients; 3) right lower lobe improved in 7 (38.9%) patients and progressed in 7 (38.9%) patients; 4) left upper lobe improved in 5 (27.8%) patients and progressed in 3 (16.7%) patients; and 5) left lower lobe improved in 8 (44.4%) patients and progressed in 4 (22.2%) patients. However, after obtaining negative RT-PCR results for viral nucleic acid, different extents of inflammatory exudation still existed in the lungs, and there was an obvious tendency of improvement, manifesting as a reduced extent and attenuation of lesions and the appearance of fibrosis (Figure S1), except for two patients with progression of the CT presentation who were demonstrated to have re-positive RT-PCR results after discharge.

**Table 4.**
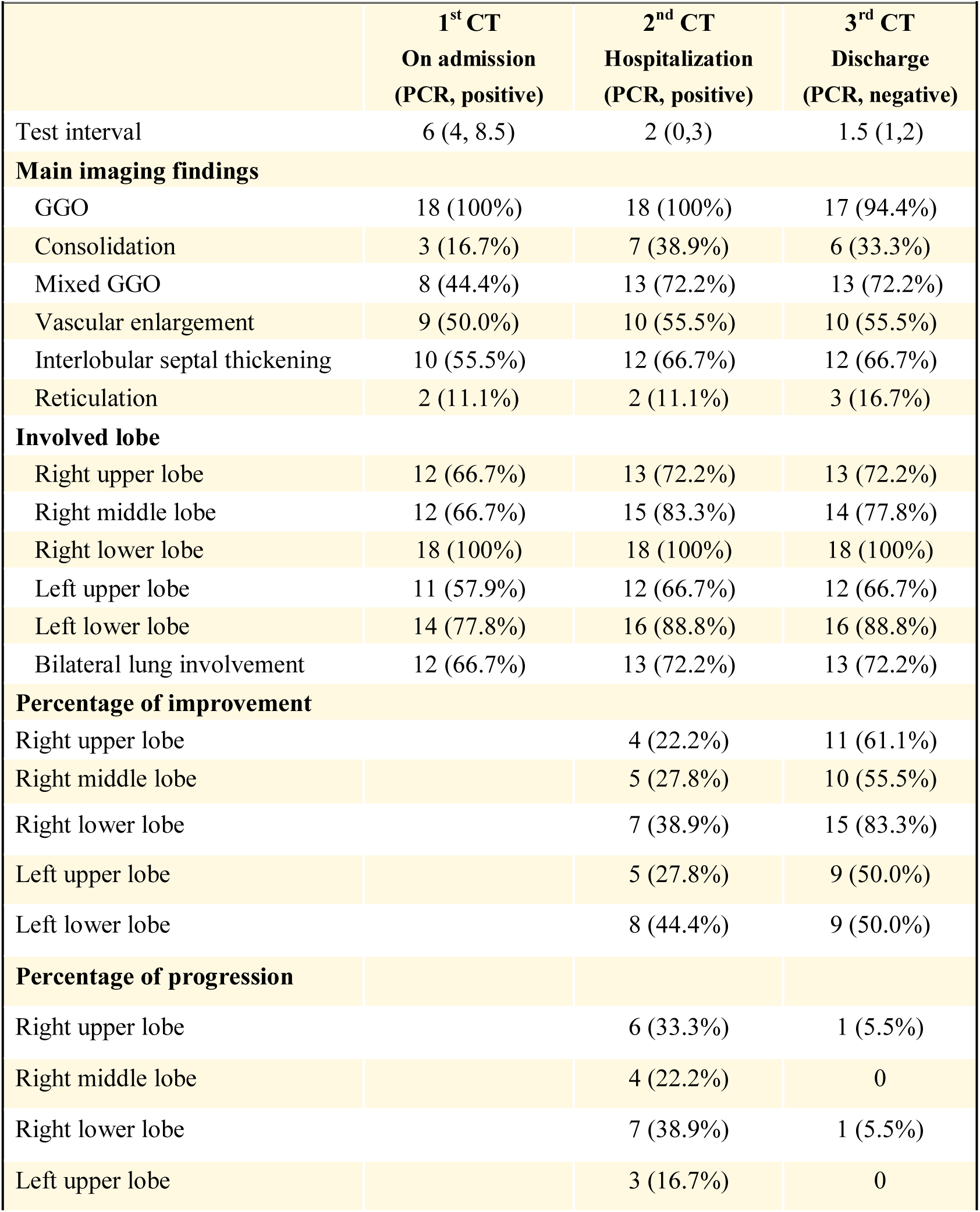
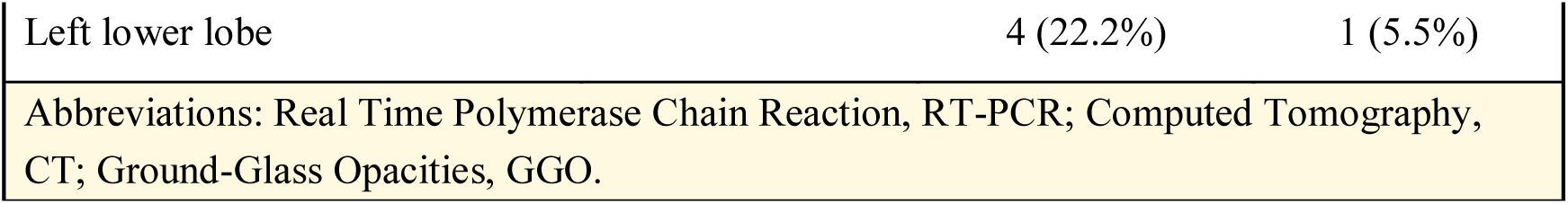
Follow-up chest CT at admission, before and after SARS-CoV-2 being negative

### Dynamic CT changes in re-admitted patients with re-positive RT-PCR results

After a median recovery of 13 days (ranging from 9 to 17 days), 7 patients had repeated positive RT-PCR results and were re-admitted to the hospital. The dynamic CT characteristics of these re-positive patients are listed in Table S1 (Supplementary appendix). Regarding the 3^rd^ CT findings with negative RT-PCR results at discharge, improvement was observed in 4 patients (case 2, case 4, case 6 and case 7, Figure S3), and progression was observed in 2 patients (case 3, Figure 1; case 5, Figure S2). Regarding the chest CT findings of patients with re-positive RT-PCR results at the time of readmission to the hospital, new GGO was present in 2 patients (case 1 and case 3 in Figure 1). The other patients improved greatly (Figure S3). In addition, after re-admission, CRP levels increased in 2 patients (case 1 and case 2), and the lymphocyte count decreased in case 1 (Figure 2). Until now, no human-to-human transmission has been found in these 7 COVID-19 patients with re-positive RT-PCR results.

**Figure 1.**
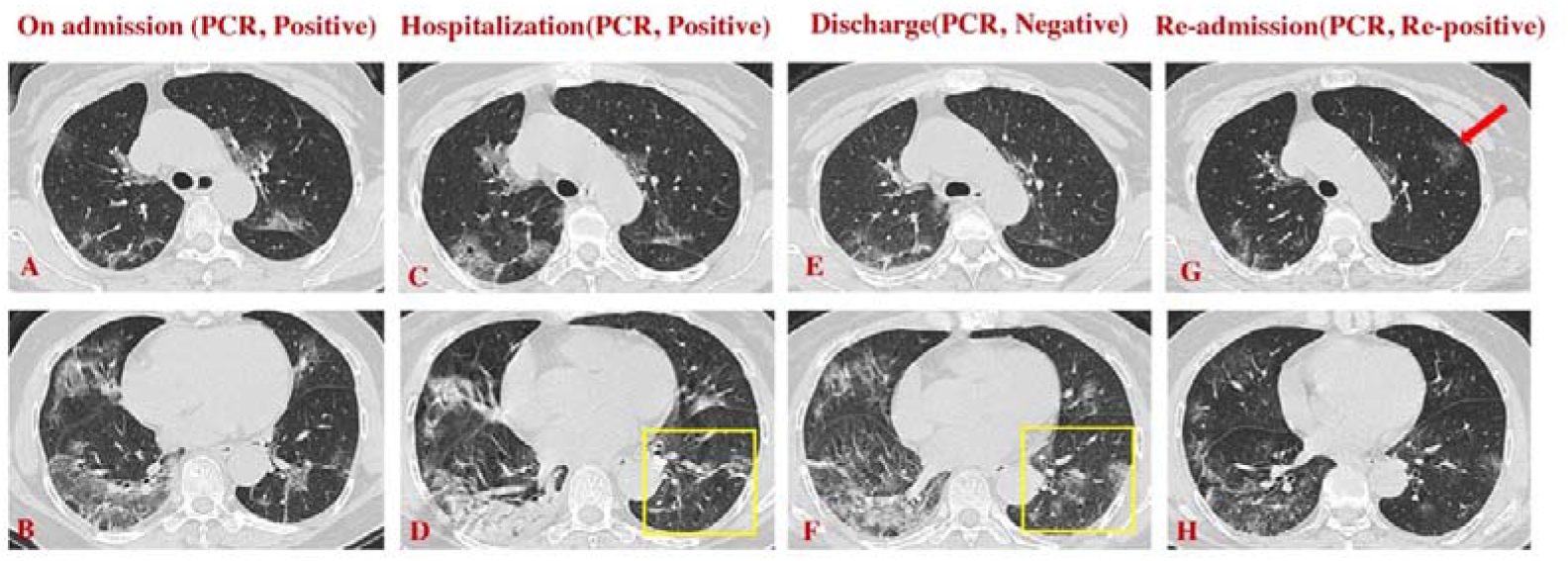
COVID-19 patient (female, 63 years old, critically severe type) who obtained positive RT-PCR results 10 days after discharge. A-B. First chest CT on admission (PCR, positive) shows a patchy area of GGO and mixed GGO in both lungs. C-D. Second chest CT in the hospital (PCR, positive) shows that the lesions had progressed and become more consolidated; E and F. Third chest CT with negative RT-PCR results (PCR, negative) shows improvement in the upper lobes and right lower lobe and progression in the left lower lobe with increasing GGO (yellow square box in F) compared with the second chest CT (yellow square box in D); G and H. Chest CT after re-admission (PCR, re-positive) shows new GGO (red arrow in G) in the left upper lobe despite great improvement in other lobes compared with the chest CT at the time of negative RT-PCR results (E).

**Figure 2.**
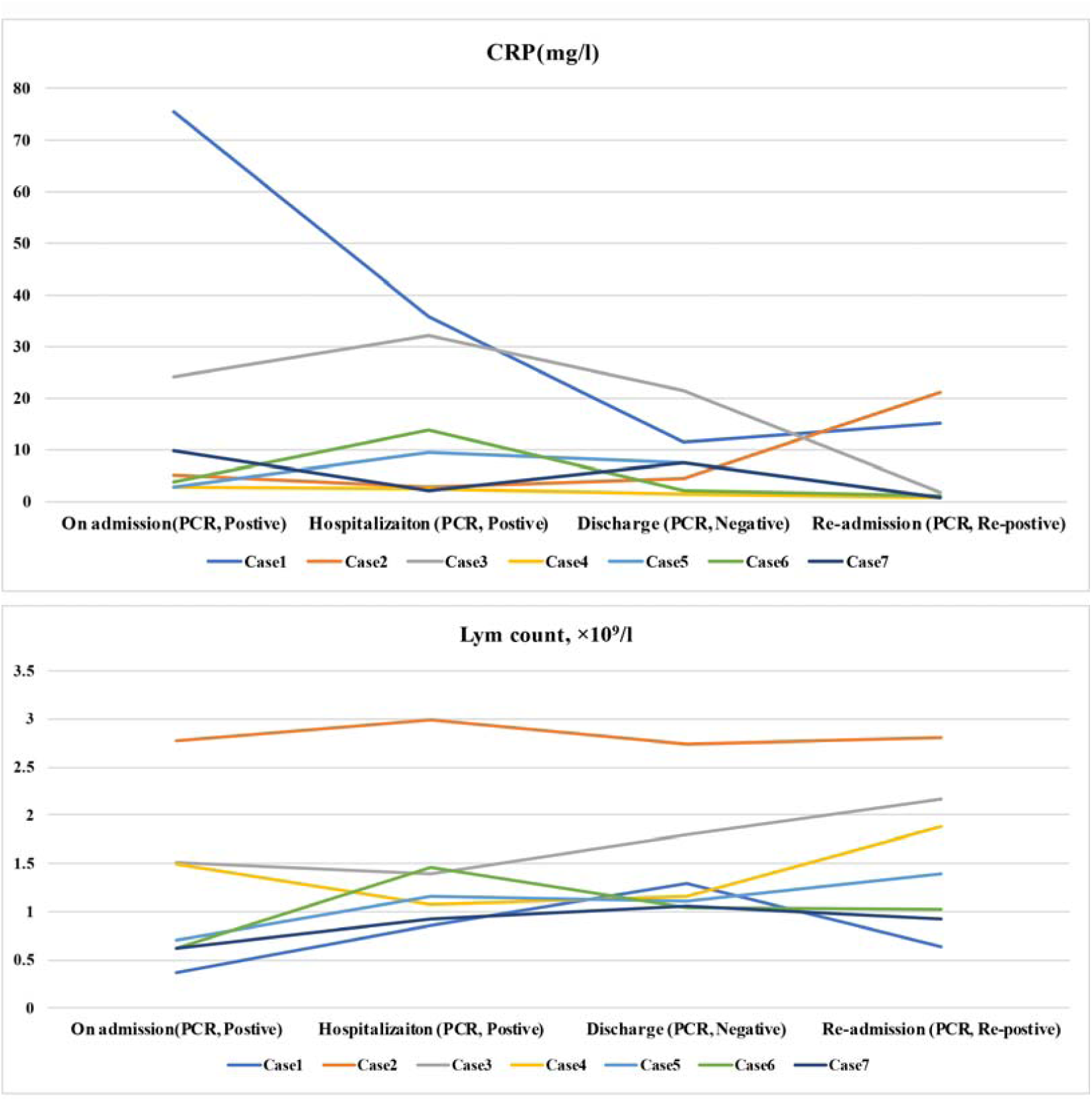
Dynamic change of CRP and lymphocyte count in the patients with re-positive RT-PCR.

## Discussion

This study analysed the dynamic variations in clinical features, laboratory tests, and CT images as well as their association with RT-PCR results. Notably, we further investigated a case series of patients who had re-positive RT-PCR results for SARS-CoV-2 nucleic acid after recovery from COVID-19 with essential management. In this research, we found the following results. 1) CRP levels and lymphocyte counts were the main significant laboratory indexes in COVID-19, 2) After obtaining negative RT-PCR results, the CRP level continuously decreased, but half of the patients still had levels higher than the normal range. 3) The lymphocyte counts of most patients increased obviously and usually recovered to the normal range after obtaining negative RT-PCR results, 4) Among our 52 patients, GGO in 47 patients (90.4%) was the most common finding on chest CT and frequently occurred in the bilateral lungs and in multi-lobular infections. Most importantly, after obtaining negative RT-PCR results, 2/18 (11.1%) patients demonstrated segmental progression on CT, and their RT-PCR results were positive again after discharge (9 days and 10 days). 5) Of 52 discharged patients, 7 patients had re-positive RT-PCR results 9 to 17 days after discharge, of whom two were found to have new GGO, and the remaining 5 patients improved compared with the CT findings with negative RT-PCR results. Thus, follow-up RT-PCR and chest CT are necessary in COVID-19, especially in recovered patients.

Human coronavirus mainly affects the respiratory system and causes mild to moderate respiratory symptoms (i.e., HCoV-229E, HCoVOC43, HCoV-NL63, and HCoV-HKU) or severe acute respiratory syndrome (SARS-CoV and MERS-CoV). Although the sequence of 2019-nCoV is relatively different from that of two other highly pathogenic coronaviruses (SARS-CoV and MERS-CoV) that threatened public health in 2003 and 2012[14,15], they may share similarities regarding pathogenesis. Chemokines and cytokines might play a key role in human coronavirus infection. Indirect evidence shows that in the second phase of SARS-CoV infection, high fever, obvious progression of pneumonia and hypoxemia occur despite a significant decline in virus titres [16]. In addition, previous research on SARS-CoV and MERS-CoV directly revealed that pro-inflammatory cytokines (IL-1, IL-6, IL12, TGFβ and IFN-γ) and chemokines (CCL2, CXCL10, and IL8) in serum increased considerably, and anti-inflammatory cytokines (IL-10) decreased in severe patients[17-24]. A similar phenomenon was observed, in which pro-inflammatory cytokines in COVID-19 increased and ICU patients had much higher plasma concentrations [6]. Cytokine storms and immunopathology via the dysregulation of chemokines and cytokines may cause epithelial and endothelial cell apoptosis, vascular leakage, and infiltration of macrophages and neutrophils, which induce severe lung injuries [25,26]. Similarly, our study demonstrated that a few COVID-19 patients had sustainable increases in CRP levels and decreases in lymphocyte counts, and most patients’ CT images demonstrated progression before the RT-PCR results became negative, which suggests that these patients may suffer from inflammatory storms and lymphatic system (especially T lymphocyte) injuries caused by the virus[7]. In addition, the serum levels of these chemokines and cytokines might remain elevated and lead to an inflammatory response even with negative RT-PCR results for SARS-CoV-2[17], which may be the reason for the progression of radiographical signs in 2 of our patients (Figure 1 and Figure S2) and the fact that almost half of the patients still had higher CRP levels after obtaining negative RT-PCR results.

Our evidence and that of previous studies indicate that a period of time is needed to reduce the inflammatory response. Research on SARS-CoV has demonstrated that after 60 days of recovery from SARS-CoV, inflammatory factors are still higher than in normal controls, pneumonia still exists in the lungs, and complete resolution may not occur until 90 days or later [17]. Thus, for COVID-19 patients discharged from the hospital, regular follow-up with laboratory tests and CT imaging are essential for monitoring inflammation progression and reduction, and sufficient nutrition support is necessary to help repair inflammatory injury. Previous radiographic studies have shown that the majority of COVID-19 patients have similar CT characteristics, such as GGO, consolidation, and fibrosis [9-11]. In improved patients, fibrosis or resolution of exudation is detected; however, new lesions or additional consolidated primary lesions are found in progressed patients.

Our research found that 7 patients had re-positive RT-PCR results 9 to 17 days after discharge. In these 7 patients, the chest CT results of 2 re-positive patients showed that the lung lesions had deteriorated compared with the chest CT results before obtaining negative RT-PCR results; one of these patients had new lesions in the lung on follow-up chest CT when readmitted to the hospital due to re-positive RT-PCR results. Our results demonstrate that even with negative RT-PCR results from nasopharyngeal swabs, patients with lesion progression on CT images should be given more attention. This phenomenon may be due to two reasons: one is that a proportion of recovered patients may still carry the virus; thus, close observation and quarantine is needed for two weeks after discharge [12]. Based on these considerations, isolation for 14 days has been added to the 6th guidelines issued by China’s National Health Commission [4]. Another reason may be that all our patients underwent nasopharyngeal swabs to test for SARS-CoV-2, which may have provided false-negative results due to sampling bias. Therefore, two rounds of negative RT-PCR results should not be recommended as the only criteria for COVID-19 recovery; the combination with dynamic chest CT is essential, as mentioned in the sixth edition of the COVID-19 patient management guidelines [4]. Additionally, for these patients with negative RT-PCR results but CT progression, deep sampling, such as lower respiratory tract or faecal sampling, is required to ensure that negative RT-PCR results are accurate. According to our research and the latest literature[27], chest CT plays a critical role in the evaluation of NCP progression as well as follow-up after RT-PCR results become negative; thus, chest CT findings should be considered as part of the standard criteria for discharge and management. In the future, more effort should be made to investigate virus retention, CT outcomes and the clinical prognosis of COVID-19.

This study has some limitations. First, we had a relatively small number of recovered patients, including only 18 of 52 patients with three follow-up CT images. Second, the time of follow-up was relatively short, and the full outcome of the RT-PCR results may be limited to some extent. Third, the results of pro-inflammatory cytokines (IL-1, IL-6, IL12, TGFβ and IFN-γ) and chemokines (CCL2, CXCL10, and IL8) were lacking, and more evidence is needed regarding the sustainable inflammatory condition of COVID-19 after obtaining negative RT-PCR results. Thus, further investigations considering the abovementioned limitations are required.

## Conclusion

Heterogeneity between CT features and RT-PCR results was found in COVID-19, especially in some recovered patients who obtained negative RT-PCR results. Our study highlights that RT-PCR and chest CT should be considered the key determinants in the clinical diagnosis and management of COVID-19 patients, such as the discharge criteria. Furthermore, long-term follow-up studies are essential in COVID-19 patients after obtaining negative RT-PCR results, especially in patients with sustainable abnormal indexes and with progression of chest CT manifestations.

## Data Availability

contact me by email

## Founding

This work was supported by 2020 Novel coronavirus pneumonia prevention and control technology project of Chengdu (NO. 2020-YF05-00007-SN); Clinical research finding of Chinese society of cardiovascular disease(CSC) of 2019 (HFCSC2019B01); the National Natural Science Foundation of China (No. 81771887, 81771897, 81971586,81901712); the Program for Young Scholars and Innovative Research Team in Sichuan Province (No. 2017TD0005) of China; and 1·3·5 project for disciplines of excellence, West China Hospital, Sichuan University (No.ZYGD18013).

